# High prevalence of soil-transmitted helminth co-infections in persons with tuberculosis in South India

**DOI:** 10.64898/2026.05.26.26353735

**Authors:** Prakash Babu Narasimhan, Komal Jain, Nonika Rajkumari, Madolyn R Dauphinais, R Jasemine, Sana Shaikh, Jainish Uresh Patel, Senbagavalli Prakash Babu, Chelsie Cintron, Meagan Karoly, Madeline E Carwile, Anne F Liu, Kimberly Maloomian, Lindsey M Locks, Saurabh Mehta, Sonali Sarkar, Urvashi B Singh, Jerrold J. Ellner, Padmini Salgame, Scott K Heysell, Natasha S. Hochberg, Subitha Lakshminarayanan, Pranay Sinha

**Affiliations:** Department of Clinical Immunology, Jawaharlal Institute of Postgraduate Medical Education and Research, Puducherry, India; Department of Preventive and Social Medicine, Jawaharlal Institute of Postgraduate Medical Education and Research, Puducherry, India; Department of Microbiology, Jawaharlal Institute of Postgraduate Medical Education and Research, Puducherry, India; University of Massachusetts Chan Medical School, Worcester, Massachusetts, USA; Boston Medical Center, Boston, Massachusetts, USA; Department of Epidemiology, Brown University School of Public Health, Providence, Rhode Island, USA; Department of Gastroenterology, Hepatology, and Endoscopy, Brigham and Women’s Hospital, Boston, Massachusetts, USA; HMX, Harvard Medical School, Boston, Massachusetts, USA; Department of Health Sciences, Boston University College of Health and Rehabilitation Sciences: Sargent College, Boston, Massachusetts, USA; Cornell Joan Klein Jacobs Center for Precision Nutrition and Health, Cornell University, Ithaca, New York, USA; Department of Microbiology, All India Institute of Medical Sciences, New Delhi, India; Center for Emerging Pathogens, Department of Medicine, New Jersey Medical School, Rutgers Biomedical and Health Sciences, Newark, New Jersey, USA; Division of Infectious Diseases and International Health, University of Virginia, Charlottesville, Virginia, USA; Department of Epidemiology, Boston University School of Public Health, Boston, Massachusetts, USA; Section of Infectious Diseases, Department of Medicine, Boston University Chobanian & Avedisian School of Medicine, Boston, Massachusetts, USA

## Abstract

Soil-transmitted helminths (STH) are a plausible but under-characterized comorbidity in tuberculosis. In this prospective South Indian cohort, multiplex stool PCR detected STH in 43% of 137 adults with pulmonary tuberculosis and 34% of 230 household contacts. Food insecurity independently predicted co-infection. Current adult deworming gaps warrant evaluation.

## Introduction

The burden of tuberculosis is disproportionately high in tropical and subtropical regions where soil-transmitted helminths (STH) are endemic.^1^ Given that tuberculosis and STH infections both concentrate in the bottom economic quintiles, high co-infection rates are likely. STH co-infections may influence tuberculosis disease progression and treatment outcomes through multiple biologically plausible mechanisms, including skewed immune signaling unfavorable for anti-tuberculosis response, malnutrition, and even impaired treatment through malabsorption of drugs and reduced adherence due to gastrointestinal symptoms.^1^

The co-prevalence of STH infections in persons with tuberculosis (PWTB) is not well-characterized. A recent systematic review included studies from additional African and Asian countries and found a pooled prevalence of approximately 29%.^2^ India accounts for a quarter of the global tuberculosis burden, but the rate of STH co-infection among persons with tuberculosis has not been well characterized.^3^ One study reported a 27.1% prevalence of STH co-infections among PWTB in Odisha but relied solely on stool microscopy, which is less sensitive than molecular assays.^4,5^

To generate rigorous, policy-relevant estimates of common intestinal STH co-infection among PWTB in India, we conducted a prospective cohort study using multiplex polymerase chain reaction–based stool diagnostics.

## Methods

### Study Design and Population

The Learning Effect of Parasites on Reinforcing Diets on Tuberculosis (TB-LEOPARD) study (NCT04584425) is a prospective cohort study conducted between April 2022 and June 2024 in Puducherry and nearby districts of Tamil Nadu (Cuddalore, Villupuram, and Tiruvannamalai). We enrolled adults (≥18 years) with newly diagnosed drug-susceptible pulmonary tuberculosis confirmed by culture or nucleic acid amplification testing through India’s National Tuberculosis Elimination Program. TB-LEOPARD recruited from the same health facilities and geographic catchment area as the Tuberculosis Learning the Impact of Nutrition (TB-LION) study, a parallel prospective study that enrolled interferon-gamma release assay-positive household contacts (HHCs) of PWTB between January 2021 and February 2024 and has been described previously.^6^ For the current analysis, we included all PWTB enrolled in TB-LEOPARD alongside HHCs enrolled in TB-LION to provide a descriptive comparison of STH prevalence across these two groups.

In the TB-LEOPARD study, we excluded individuals with prior tuberculosis treatment, drug-resistant tuberculosis, inability to provide a stool sample, or severe comorbidities. Because TB-LION provided nutritional support, we excluded persons with extreme undernutrition (BMI <14 kg/m^2^), bilateral leg edema, or severe peripheral neuropathy due to increased risk of refeeding syndrome and Wernicke’s encephalopathy. We excluded pregnant persons because pregnancy-related changes in weight would confound longitudinal assessment of weight gain. Institutional review boards at the Jawaharlal Institute of Postgraduate Medical Education and Research (JIPMER) and the Boston University Medical Campus approved the study. All participants provided written informed consent in Tamil.

### Data Collection and Laboratory Methods

At enrolment, trained study staff collected baseline data using standardized questionnaires to capture sociodemographic characteristics, tobacco use, alcohol misuse (AUDIT-C), household food insecurity (Household Food Insecurity Access Scale), and multidimensional poverty (Oxford Multidimensional Poverty Index).^7,8^ Study staff also measured anthropometric indicators, including weight, height, mid-upper arm circumference, and triceps and subscapular skinfold thickness.^9^

Participants provided a fresh stool sample at baseline. Study staff transported samples on ice to the JIPMER laboratory, where technicians sonicated the stool and extracted DNA from approximately 200 mg of specimen. We used a multiplex real-time polymerase chain reaction (PCR) assay to detect common intestinal STH, including major STH (*Ancylostoma duodenale, Necator americanus, Ascaris lumbricoides, Strongyloides stercoralis*, and *Trichuris trichiura*) using published primer–probe sets with appropriate controls.^10^ We defined positivity as amplification with a cycle threshold ≤36. For household contacts enrolled in TB-LION, laboratory staff analyzed stool specimens using the same multiplex PCR platform and primer–probe sets; however, samples were transported in zinc polyvinyl alcohol and 10% formalin and were not sonicated prior to DNA extraction.

### Outcomes and Statistical Analysis

The primary outcome was the presence of any STH co-infection detected by multiplex stool PCR. To contextualize helminth prevalence observed among persons with tuberculosis, we descriptively compared the prevalence of PCR-detected STH infection in TB-LEOPARD participants with that among HHCs enrolled in TB-LION using the χ^2^ test. To identify factors independently associated with STH co-infection, we used multivariable logistic regression in the combined sample of PWTB and HHCs, estimating ORs and 95% confidence intervals (CIs). We included age, sex, and participant type (PWTB vs. HHC) as a priori covariates, and additionally included covariates with p<0.20 in univariable analysis. Alcohol misuse and smoking were retained in the full adjusted model given their a priori biological plausibility as confounders, even after attenuation in multivariable analysis. We also generated maps to illustrate enrolment coverage and to assess whether STH infections clustered geographically.

## Results

Among 137 PWTB, the mean age was 35.4 years (SD 13.2) and 42 (30.7%) were female.Among 230 HHCs, the mean age was 33.2 years (SD 10.8) and 143 (62.2%) were female. PWTB had substantially higher rates of severe undernutrition (BMI <16.0 kg/m^2^: 36.5% vs 3.5%), alcohol misuse (31.4% vs 4.3%), and tobacco use (31.4% vs 3.9%) compared to HHCs (all p<0.001). Food insecurity was more prevalent among PWTB (27.0% vs 17.6%; p=0.035), while employment rates were lower (25.5% vs 51.7%; p<0.001). Poverty did not differ significantly between groups (Supplementary Table 2).

Among the total cohort of 367 participants, 138 (37.6%) had at least one STH detected by PCR. Prevalence was higher among PWTB (59/137, 43.1%) than among HHCs (79/230,34.3%), though this difference was not statistically significant (p=0.096; Supplementary Table 1). *Necator americanus* was the most frequently identified species overall (n=141, 38.4%), followed by *Strongyloides stercoralis* (n=47, 12.8%); this pattern was consistent across both groups (Supplementary Table 1). Among the 138 participants with PCR-confirmed STH infections, most had single-species infections; multiple STH infections were present in 12 PWTB (20.3%) and a corresponding proportion of HHCs, with *S. stercoralis* and *N. americanus* being the most common co-infecting pair. Infections with *Ancylostoma duodenale* (n=22) and *Trichuris trichiura* (n=11, all among HHCs) were less common (Figure 1). Helminth infections were detected in participants enrolled in every calendar month (Supplementary Figure 1), and no geographic clustering or restriction to rural areas was evident (Supplementary Figure 2).

**Figure 1:**
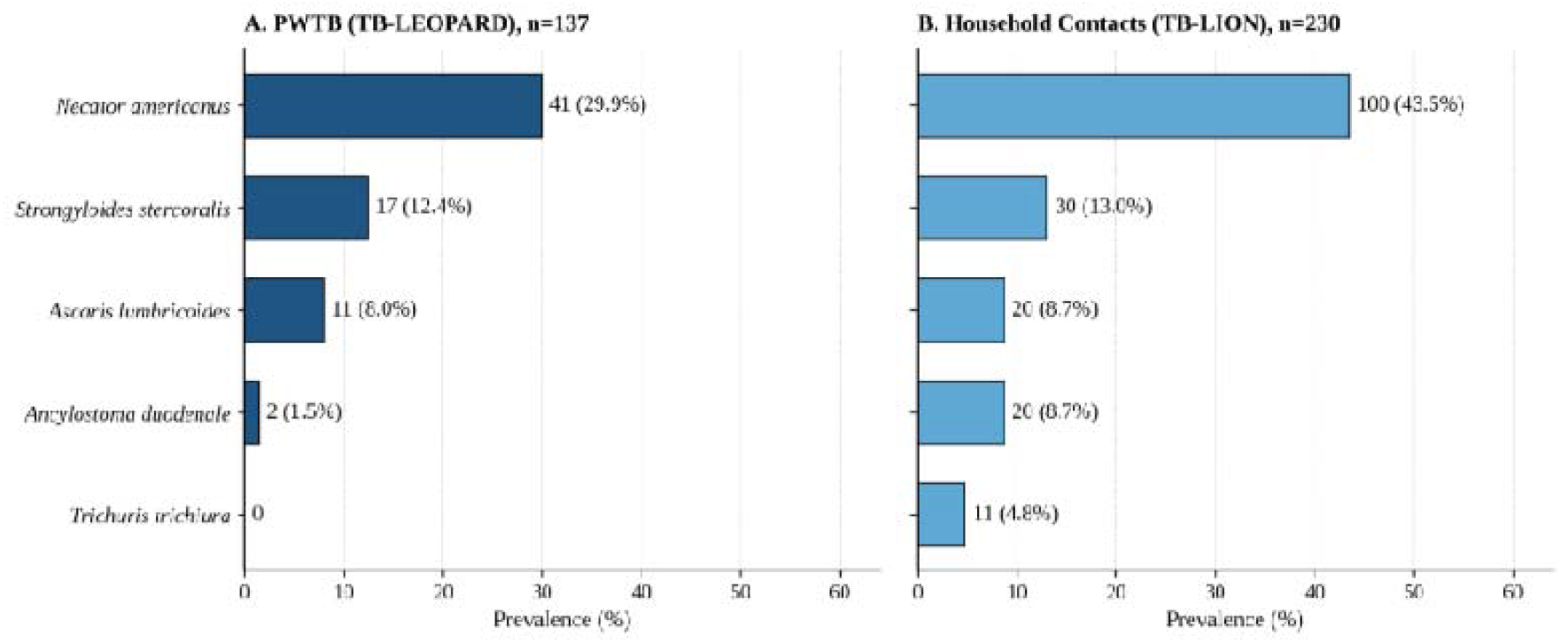
Distribution of STH species among adults with pulmonary tuberculosis (n=137) and household contacts (n=230) in South India

Gastrointestinal symptoms were uncommon and did not differ significantly between participants with and without STH co-infection. Diarrhea, abdominal pain, bloating, nausea, and vomiting were each reported in fewer than 5% of participants in either group (Supplementary Table 4).

In the combined cohort, participants with STH co-infection were more likely to be older, food insecure, and to report alcohol misuse or smoking. Rates of severe undernutrition (BMI <16.0 kg/m^2^) were higher among those with STH co-infection (20.3%) than those without (13.1%). Employment status and poverty did not differ meaningfully between groups (Supplementary Table 3).

In multivariable logistic regression, food insecurity remained independently associated with STH co-infection after adjusting for age, sex, participant type, alcohol misuse, smoking, employment, and poverty (adjusted OR 2.10; 95% CI 1.22–3.62; p=0.007). Older age was also independently associated with STH co-infection (adjusted OR per year 1.03; 95% CI 1.01–1.05; p=0.016). Severe undernutrition, alcohol misuse, and smoking were each associated with STH co-infection in univariable analyses but were attenuated after full adjustment. Participant type, sex, employment, and poverty were not independently associated (Table 1). We found no evidence of effect modification by sex, smoking, or alcohol misuse (all p>0.15). The interaction term for age group (p=0.055) did not reach statistical significance and should be interpreted cautiously; stratum-specific estimates are shown in Supplementary Table 5.

**Table 1:** Univariable and Multivariable Logistic Regression for Factors Associated with Soil-Transmitted Helminth Co-infection (N=367)

| Variable | Crude OR (95% CI) | P-value | Full Adjusted aOR (95% CI) | P-value |
| --- | --- | --- | --- | --- |
| **Participant Type (PWTB vs. HHC)** | 1.45 (0.94–2.23) | 0.096 | 0.88 (0.48–1.61) | 0.682 |
| **Age (per year increase)** | 1.03 (1.01–1.05) | **<0.001** | 1.03 (1.01–1.05) | **0.016** |
| **Sex (Male vs. Female)** | 1.36 (0.89–2.07) | 0.158 | 1.08 (0.64–1.83) | 0.761 |
| **BMI Category (kg/m<sup>2</sup>)** |  |  |  |  |
| ≥18.5 (Normal/Overweight) | Reference | — | Reference | — |
| 16.0–18.5 (Underweight) | 1.29 (0.79–2.11) | 0.306 | 1.21 (0.69–2.12) | 0.510 |
| <16.0 (Severely Underweight) | 1.84 (1.02–3.32) | **0.043** | 1.39 (0.66–2.92) | 0.386 |
| **Food Insecurity (Insecure vs. Secure)** | 2.31 (1.38–3.86) | **0.002** | 2.10 (1.22–3.62) | **0.007** |
| **Alcohol Misuse (Yes vs. No)** | 2.49 (1.38–4.49) | **0.002** | 1.30 (0.55–3.05) | 0.547 |
| **Smoking Status (Ever vs. Never)** | 2.38 (1.32–4.32) | **0.004** | 1.37 (0.61–3.12) | 0.448 |
| **Employment Status (Employed vs. Unemployed)** | 1.16 (0.76–1.78) | 0.499 | 1.05 (0.65–1.72) | 0.833 |
| **Below Poverty Line (<Rs. 10,000/mo)** | 0.54 (0.19–1.52) | 0.243 | 0.58 (0.19–1.75) | 0.332 |

## Discussion

To our knowledge, this is the first study in India to estimate the prevalence of tuberculosis and STH co-infection using molecular diagnostics in TB-affected households. PWTB had high rates of largely asymptomatic STH infection, with higher prevalence among PWTB than HHCs. Importantly, infections in PWTB were detected among both urban and rural participants and throughout the year, indicating that STH co-infection among PWTB in South India is not confined to rural settings or seasonal exposure. Hookworms, the most common species detected, are a well-established cause of iron deficiency anemia, a recognized risk factor for adverse tuberculosis outcomes and mortality. Approximately one in five STH-infected PWTB harbored multiple STH species despite minimal gastrointestinal symptoms, suggesting that STH infections may represent a largely silent but important contributor to nutritional and immunologic stress among PWTB.

In the TB-LEOPARD cohort, more than three-quarters of participants experienced multidimensional poverty. Household food insecurity, which is often exacerbated by the catastrophic costs of tuberculosis, remained the strongest predictor of STH co-infection.^11^ This study was not designed to assess whether parasitic infections increase susceptibility to tuberculosis or disease progression; rather, our findings underscore the clustering of tuberculosis and intestinal parasitic infections within the same structurally vulnerable populations, shaped by shared socioeconomic and environmental risks.

Notably, routine bi-annual deworming programs are already implemented in our study setting, raising questions about the adequacy of current strategies for reducing STH burden among adults. In India, mass deworming efforts have largely focused on children and pregnant women, while non-pregnant adults—who comprise the majority of individuals affected by tuberculosis—are not routinely targeted. Our results suggest that this programmatic gap may allow STH infection to persist among adults with tuberculosis despite ongoing population-level deworming efforts.

Our study has limitations. It was conducted in a single region with a modest sample size, which limits precision and generalizability to other Indian ecologies. As we excluded individuals with very severe undernutrition to avoid refeeding syndrome and Wernicke’s encephalopathy, we may have attenuated anthropometric differences between those with and without STHs. Differences in stool processing and in demographic characteristics between PWTB and HHCs may have resulted in higher nucleic acid yields in the latter.Crucially, molecular assays are highly sensitive but may detect residual DNA from prior infections, leading to possible non-differential misclassification. Unmeasured confounding, including household sanitation and environmental exposures, may remain.

Despite these caveats, STH infections remain a common and under-addressed comorbidity in households affected by tuberculosis. Our findings raise the question of whether empiric deworming of PWTB and HHCs at treatment initiation warrants prospective evaluation; randomized studies are needed to determine whether treating STH co-infections improves tuberculosis treatment outcomes. Nutritional support with counseling on food and water safety and footwear use may offer complementary benefit and is consistent with existing guidelines.^12^ Future studies should assess the impact of such interventions on tuberculosis outcomes and nutritional recovery.

Ultimately, durable reductions in STH burden will likely require attention to upstream determinants, including food insecurity, sanitation, and living conditions, underscoring the need for integrated social and public health approaches alongside biomedical care.

## Data Availability

All data produced in the present study are available upon reasonable request to the authors.

