## Supplementary Material for "High prevalence of soil-transmitted helminth co-infections in persons with tuberculosis in South India"

**Supplementary Figure 1: Monthly Prevalence of Soil-Transmitted Helminth Infection in Persons with Tuberculosis and Household Contacts**

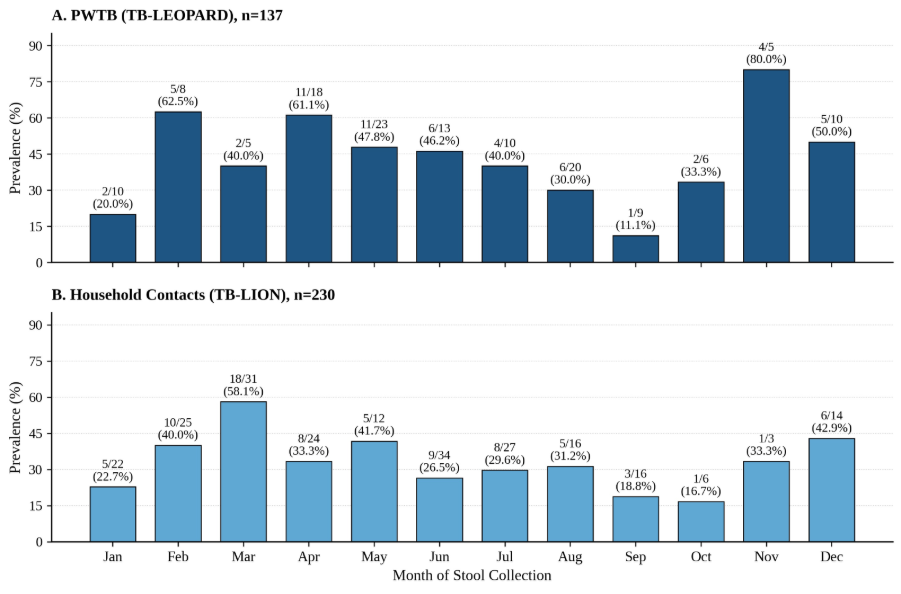

**Supplementary Figure 2:** Spatial distribution of study participants by soil-transmitted helminth co-infection status, shown separately for (A) persons with pulmonary tuberculosis enrolled in TB-LEOPARD (n=137) and (B) IGRA-positive household contacts enrolled in TB-LION (n=230). Markers indicate participant locations across Tamil Nadu districts, color-coded by STH co-infection status. Maps illustrate enrollment coverage and were generated to assess whether STH infections clustered geographically.

**A.**
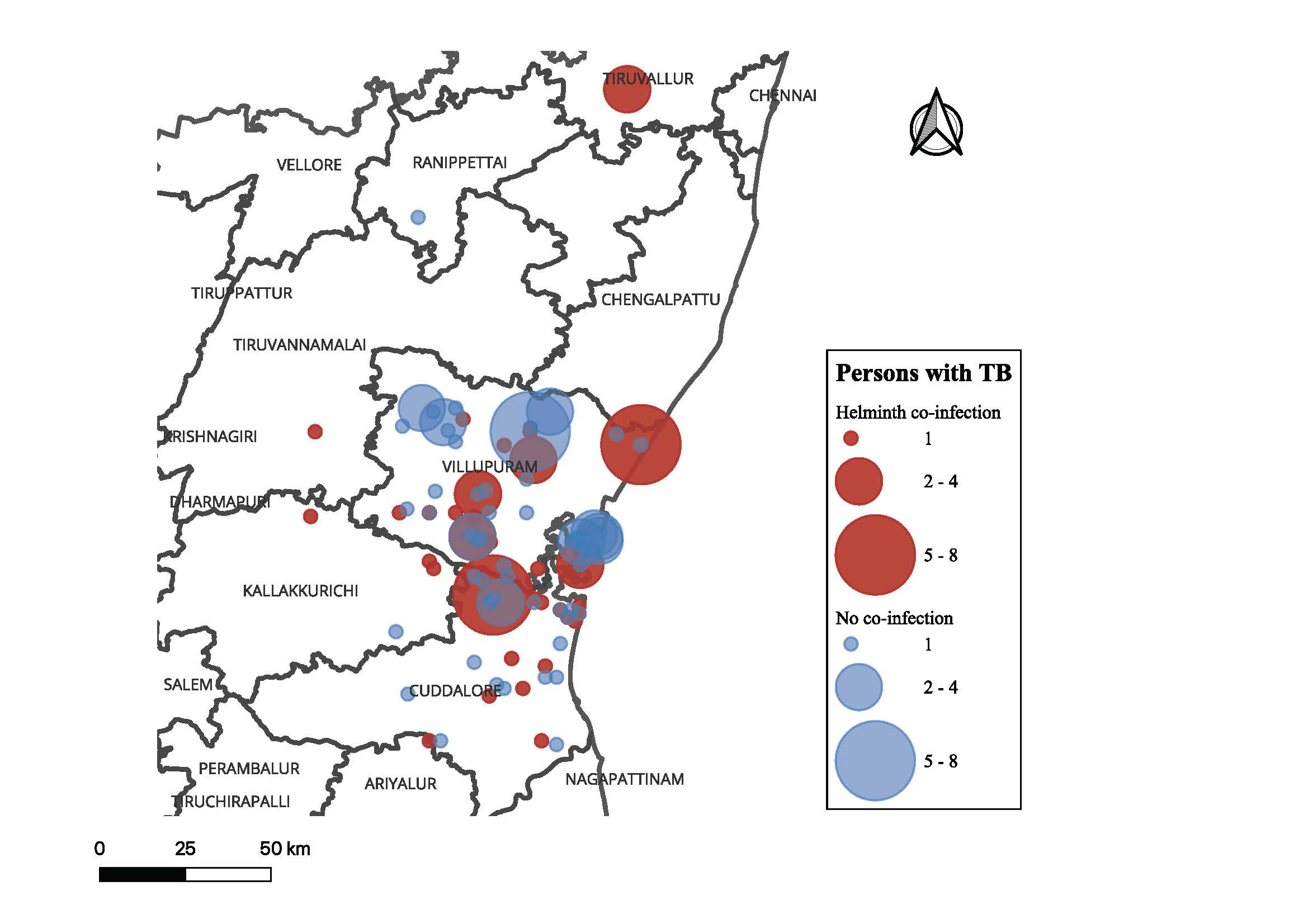

**B.**
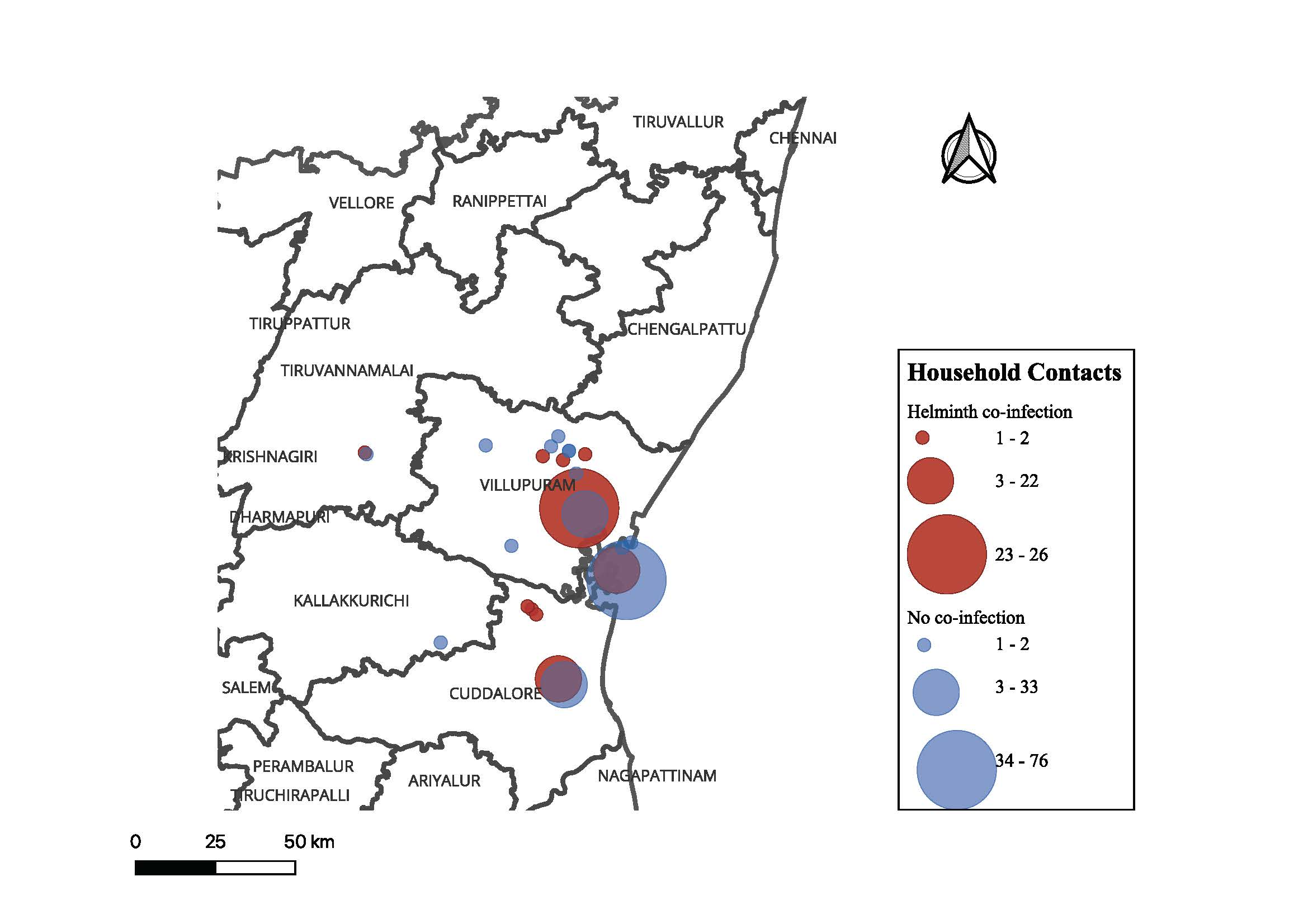

**Supplementary Table 1:** Soil-Transmitted Helminth Species Prevalence Between Persons with Active Tuberculosis (n=137) and IGRA-positive Household Contacts (n=230)

| **Organism** | **PWTB (n=137)** | **HHC (n=230)** | **Total (N=367)** |
| --- | --- | --- | --- |
| **Any STH Positive** | 59 (43.1%) | 79 (34.3%) | 138 (37.6%) |
| Necator americanus | 41 (29.9%) | 100 (43.5%) | 141 (38.4%) |
| Strongyloides stercoralis | 17 (12.4%) | 30 (13.0%) | 47 (12.8%) |
| Ancylostoma duodenale | 2 (1.5%) | 20 (8.7%) | 22 (6.0%) |
| Ascaris lumbricoides | 11 (8.0%) | 20 (8.7%) | 31 (8.4%) |
| Trichuris trichiura | 0 (0.0%) | 11 (4.8%) | 11 (3.0%) |

**Supplementary Table 2:** Baseline Characteristics by Participant Type

| **Characteristic** | **PWTB (n=137)** | **HHC (n=230)** | **P-value** |
| --- | --- | --- | --- |
| **Any STH Co-infection, n/N (%)** | 59/137 (43.1%) | 79/230 (34.3%) | 0.096 |
| **Age (years), Mean (SD)** | 35.4 (13.2) | 33.2 (10.8) | 0.091 |
| **Female Sex, n/N (%)** | 42/137 (30.7%) | 143/230 (62.2%) | **<0.001** |
| **Employed, n/N (%)** | 35/137 (25.5%) | 119/230 (51.7%) | **<0.001** |
| **Below Poverty Line, n/N (%)** | 31/136 (22.8%) | 62/227 (27.3%) | 0.406 |
| **Food Insecure, n/N (%)** | 37/137 (27.0%) | 39/221 (17.6%) | **0.035** |
| **BMI Category (kg/m²), n/N (%)** |  |  | **<0.001** |
| Severely Underweight (<16.0) | 50/137 (36.5%) | 8/230 (3.5%) |  |
| Underweight (16.0–18.5) | 50/137 (36.5%) | 51/230 (22.2%) |  |
| Normal/Overweight (≥18.5) | 37/137 (27.0%) | 171/230 (74.3%) |  |
| **Alcohol Misuse, n/N (%)** | 43/137 (31.4%) | 10/230 (4.3%) | **<0.001** |
| **Former or Current Smoker, n/N (%)** | 43/137 (31.4%) | 9/230 (3.9%) | **<0.001** |

**P-values:** For continuous variables (Age), Satterthwaite or pooled t-test p-values are reported. For categorical variables including BMI category, Chi-Square p-values are reported (without continuity correction).

Supplementary Table 3: Baseline Characteristics by STH Co-infection Status

| **Characteristic** | **Parasite Negative (n=229)** | **Parasite Positive (n=138)** | **P-value** |
| --- | --- | --- | --- |
| **Participant Type, n/N (%)** |  |  |  |
| Persons with Tuberculosis | 78/229 (34.1%) | 59/138 (42.8%) | 0.096 |
| Household Contacts | 151/229 (65.9%) | 79/138 (57.2%) |  |
| **Age (years), Mean (SD)** | 32.4 (11.7) | 36.7 (11.5) | **<0.001** |
| **Female Sex, n/N (%)** | 122/229 (53.3%) | 63/138 (45.7%) | 0.157 |
| **Employed, n/N (%)** | 93/229 (40.6%) | 61/138 (44.2%) | 0.500 |
| **Below Poverty Line, n/N (%)** | 15/227 (6.7%) | 5/136 (3.7%) | 0.236 |
| **Food Insecure, n/N (%)** | 35/222 (15.8%) | 41/136 (30.1%) | **0.001** |
| **BMI Category (kg/m²), n/N (%)** |  |  | **0.038** |
| Severely Underweight (<16.0) | 30/229 (13.1%) | 28/138 (20.3%) |  |
| Underweight (16.0–18.5) | 61/229 (26.6%) | 40/138 (29.0%) |  |
| Normal/Overweight (≥18.5) | 138/229 (60.3%) | 70/138 (50.7%) |  |
| **Alcohol Misuse, n/N (%)** | 23/229 (10.0%) | 30/138 (21.7%) | **0.002** |
| **Former or Current Smoker, n/N (%)** | 23/229 (10.0%) | 29/138 (21.0%) | **0.004** |

**P-values:** For continuous variables (Age), Satterthwaite or pooled t-test p-values are reported. For categorical variables including BMI category, Chi-Square p-values are reported. *BMI category p-value calculated using Mantel-Haenszel Chi-Square test.

**Supplementary Table 4: Gastrointestinal Symptoms**

| **Gastrointestinal Symptom** | **Parasite Negative (n=229)** | **Parasite Positive (n=138)** | **P-value (Fisher’s)** |
| --- | --- | --- | --- |
| Diarrhea | 2 (0.9%) | 0 (0.0%) | 0.530 |
| Stomach Pain | 10 (4.4%) | 3 (2.2%) | 0.385 |
| Bloating | 2 (0.9%) | 2 (1.4%) | 0.633 |
| Constipation | 1 (0.4%) | 3 (2.2%) | 0.151 |
| Nausea | 9 (3.9%) | 5 (3.6%) | 1.000 |
| Vomiting | 4 (1.7%) | 2 (1.4%) | 1.000 |

**Supplementary Table 5:** Effect Modification of the Association Between Food Insecurity and Soil-Transmitted Helminth Co-infection by Selected Covariates

| **Effect Modifier** | **Stratum** | **Adjusted OR (95% CI) – Food Insecure vs. Secure** | **Interaction P-value** |
| --- | --- | --- | --- |
| **Sex** | Female | 2.84 (1.33–6.04) | 0.257 |
|  | Male | 1.51 (0.68–3.32) |  |
| **Age Group** | <45 years | 2.67 (1.43–4.99) | 0.055 |
|  | ≥45 years | 1.10 (0.39–3.15) |  |
| **Smoking Status** | Never smoker | 1.92 (1.06–3.49) | 0.484 |
|  | Ever smoker | 3.24 (0.85–12.31) |  |
| **Alcohol Misuse** | No misuse | 2.21 (1.20–4.07) | 0.721 |
|  | Misuse | 1.74 (0.54–5.62) |  |

*CI denotes confidence interval
